# The Prevalence of RT-PCR Positivity of SARS-CoV-2 on 10,000 Patients from Three Cities Located on the Eastern of Turkey

**DOI:** 10.1101/2020.06.25.20138131

**Authors:** Karamese Murat, Ozgur Didem, Tarhan Ceyda, Dik Altintas Susamber, Caliskan Okan, Tuna Aysegul, Kazci Saliha, Karadavut Mursel, Gumus Abdullah, Apaydin Gozde, Mumcu Necati, Coruh Onur, Tutuncu Emin Ediz

## Abstract

COVID-19, is caused by SARS-CoV-2, has been started on December/2019 in Wuhan/China and spread all over the world. We analyzed RT-PCR results of 10,000 cases from April-2 to May-30, 2020 in three neighbor cities located on the Eastern of Turkey. The final study population was 7853 cases after excluded screening tests. RT-PCR were performed to detect the SARS-CoV-2-specific RdRp (RNA-dependent-RNA-polymerase) gene fragment. The number of total positive samples out of 7853 were 487; however, the number of non-repeating positive patient was 373 (4.8%). The cough and fever were the most common symptoms in positive cases. The epidemiologic studies should be performed about the prevalence of SARS-CoV-2 infection to better understand the effect of the virus all over the world.

## Introduction

Coronavirus disease (COVID-19), which is caused by SARS-CoV-2, has been started on the December of 2019 at the wet animal market in Wuhan City, China and spread all over the world [1, 2]. The disease is characterized by high fever, cough, shortness of breath, pneumonia, losing the sense of smelling/tasting, and other respiratory tract symptoms, and became a great global public health concern [3]. The Real Time Polymerase Chain Reaction (RT-PCR) analysis was used to detect the SARS-CoV-2 virus from respiratory specimens such as tracheal aspirate, nasopharyngeal swab, and sputum [4]. On the other hand, some physicians argued that Computed Tomography (CT) imaging may help the identification of SARS-CoV-2 infection, even the RT-PCR results were negative [5, 6]. From now on, RT-PCR is a gold-standard diagnostic method and has been provided more accuracy and quick diagnosis for SARS-CoV-2 detection within nearly two hours [4].

In this study, we retrospectively analyzed RT-PCR results of 10,000 cases from April 2 to May 30, 2020 in Kars, Iğdır, and Ardahan cities that are located on the Eastern of Turkey. After the screening tests were excluded, the final study population was 7853 cases. All the cases were suspected of SARS-CoV-2 infection because of the symptoms or close contact with a COVID-19 patient. RT-PCR were performed to detect the SARS-CoV-2-specific RdRp (RNA-dependent RNA polymerase) gene fragment from respiratory specimens such as tracheal aspirate, nasopharyngeal swab, and sputum.

## Methods

### Data collection

The study was approved by both the Republic of Turkey Ministry of Health COVID-19 Scientific Research Evaluation Commission (Approval date: 02.05.2020; number: 2020-05-03T18_02_30) and the Local Ethics Committee of Kafkas University Faculty of Medicine (Approval date: 06.05.2020 number: 80576354-050-99/129). A total of 10,000 cases from April 2 to May 30 were tested for SARS-CoV-2 infection in Kars, Iğdır, and Ardahan cities that are located on the Eastern of Turkey; however, 7853 cases were evaluated who had typical respiratory infection symptoms such as fever, cough and shortness of breath, or close contact with a COVID-19 patient. In this study, the screening RT-PCR test results (n=2147) were excluded (Figure 1). All RT-PCR tests from three cities were performed at Kafkas University, Faculty of Medicine, Department of Medical Microbiology, Molecular Microbiology Laboratory, Kars, Turkey.

**Figure 1:**
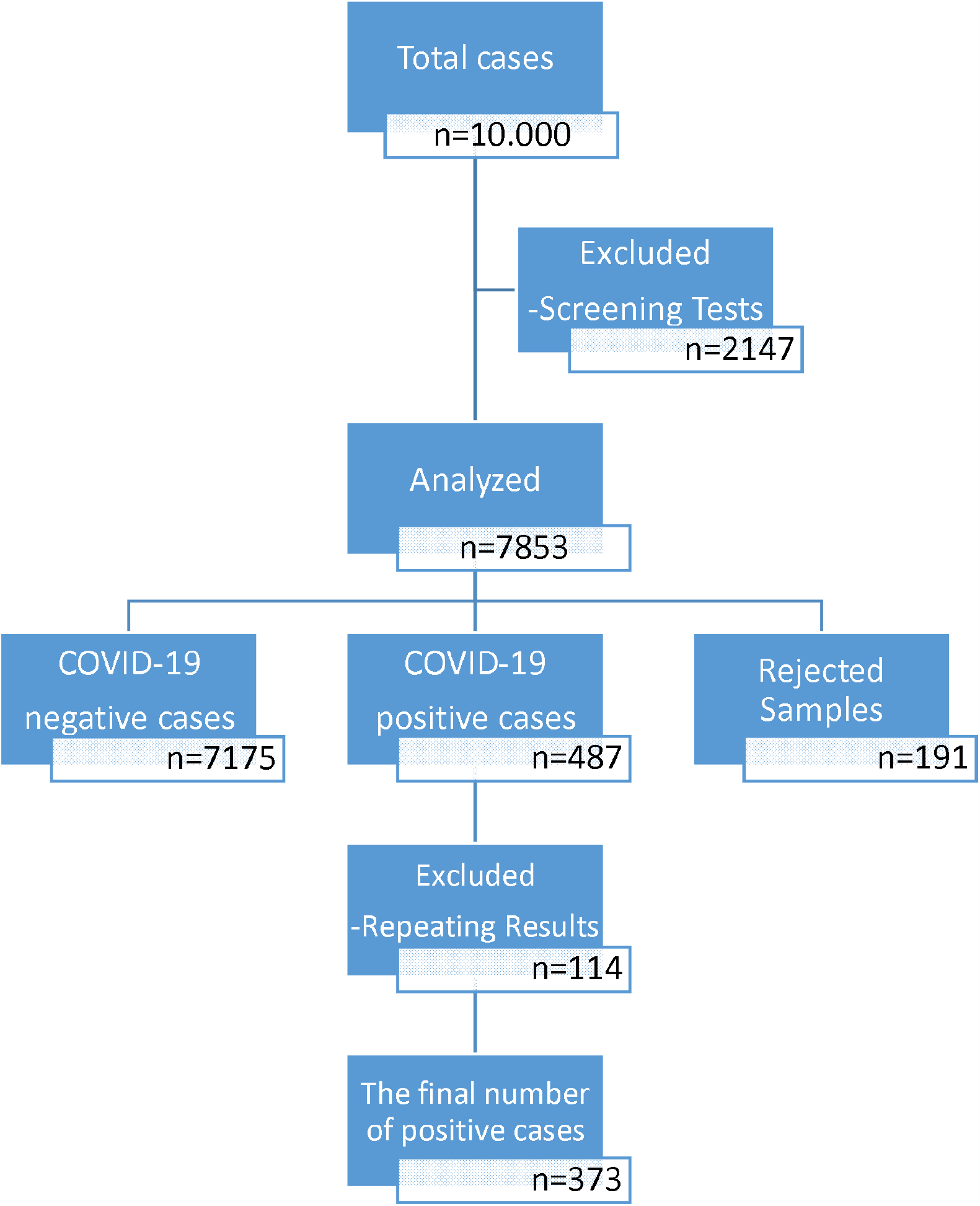
Study flow diagram.

### RT-PCR Protocol

The detection of SARS-CoV-2 virus in respiratory specimens especially in nasopharyngeal swabs was detected by Real-Time PCR technique via using Roche Lightcycler-96 device (Roche Diagnostic Systems, Basel, Switzerland). RT-PCR was performed using SARS-CoV-2 (2019-nCoV) qPCR Detection Kit (Bioeksen R&D, İstanbul, Turkey) that is targeting SARS-CoV-2-specific RdRp gene fragment. The final PCR concentration was 20-µl (10-µl qPCR master mix, 5-µl primer/probe set, and 5-µl template). The nucleic acid amplification was performed with the following PCR steps: Reverse Transcription Stage (45°C, 15 min, 1 cycle), Initial activation Stage (95°C, 3 min, 1 cycle), and Amplification Stage (Denaturation: 95°C, 5 sec, Annealing and extension: 55°C, 35 sec, 45 cycles). All samples were run together with a SARS-Cov-2 positive control, and negative control (H_2_O). For data analysis, 2-^ΔΔ^ CT method was used and CT values were less than 40 was defined as a positive test.

### Statistical Methods

Statistical analysis was performed with the Statistical Package for the Social Sciences version 22.0 (SPSS, IBM, Armonk, NY, USA). The “number,” “percentage,” “mean,” and “standard deviation” were used for the descriptive statistics of the continuous variables. The independent samples t-test or Mann-Whitney U test were used to compare two independent groups. The Pearson Chi-Square or Fisher’s exact tests were used to analyzing categorical data. The results were evaluated with a confidence interval of 95%, and the level of significance, p, was set at 0.05.

## Results

A total of 7853 cases were included from three neighbor cities. Of those 4642 (59.1%) were male, 3211 (40.8%) were female. The mean age of all cases 38.14±21.10. The nasopharyngeal swab samples (n=7826, 99.7%) were the most preferred respiratory specimens. The detailed descriptive characteristics of all patients are in Table 1. The number of total positive sample was 487; however, the number of non-repeating positive patient was 373 (4.8%). The distribution of non-repeating positive patients were as following; Kars (n=181, 48.5%), Iğdır (n=147, 39.4%), and Ardahan (n=45, 12.1%). Of those, 156 (41.8%) were male and 217 (58.2%) were female. The mean age of positive cases was 36.96±20.25. The cough and fever were the most common symptoms in positive cases (n= 111, 29.8% and n=98, 26.3%, respectively); however, most of positive cases were asymptomatic (n=198). Additionally, respiratory diseases such as asthma and chronic obstructive pulmonary disease (COPD) were the most recorded additional diseases for positive cases. The detailed information about the positive cases are in Table 2. The statistical analysis showed that there was significant difference between positive and negative cases regarding gender (**X**^**2**^**= 50**.**903, p=0**.**000**), while no statistically significant difference was detected between them concerning age (Z= −0.980, p=0.327).

**Table 1:** The descriptive characteristics of all patients from three neighbor cities

|  | Kars |  | İğdir |  | Ardahan |  | Total |
| --- | --- | --- | --- | --- | --- | --- | --- |
|  | n | % | n | % | n | % | n |
| **Number of samples** | 3509 | 44.7 | 3130 | 39.9 | 1214 | 15.4 | 7853 |
| **Gender** |  |  |  |  |  |  |  |
| *Male* | 2120 | 60.9 | 1758 | 56.2 | 764 | 62.9 | 4642 |
| *Female* | 1389 | 39.1 | 1372 | 43.8 | 450 | 37.1 | 3211 |
| **Mean age** | 38.54±21.48 |  | 35.23±19.80 |  | 44.33±21.77 |  | 38.14±21.10 |
| **Median age** | 34 |  | 31 |  | 41 |  | 33 |
| **Age range** |  |  |  |  |  |  |  |
| *0-20* | 596 |  | 575 |  | 120 |  | 1291 |
| *21-40* | 1516 |  | 1539 |  | 494 |  | 3549 |
| *41-60* | 753 |  | 637 |  | 302 |  | 1692 |
| *60+* | 644 |  | 379 |  | 298 |  | 1321 |
| **Specimen types** |  |  |  |  |  |  |  |
| *Nasopharyngeal swab* | 3506 | 99.9 | 3130 | 100 | 1190 | 98.0 | 7826 |
| *Sputum* | 0 | 0 | 0 | 0 | 24 | 2.0 | 24 |
| *Tracheal aspirate* | 3 | 0.1 | 0 | 0 | 0 | 0 | 3 |
| **Results** |  |  |  |  |  |  |  |
| *Total Positive* | 210 | 5.98 | 228 | 7.3 | 49 | 4.0 | 487 |
| *Negative* | 3243 | 92.4 | 2797 | 89.3 | 1135 | 93.8 | 7175 |
| *Rejected* | 56 | 1.6 | 105 | 3.5 | 30 | 2.5 | 191 |
| ***Non-repeating positives*** | **181** | **5.15** | **147** | **4.7** | **45** | **3.70** | **373** |

**Table 2:** The descriptive characteristics of positive patients from three neighbor cities

|  | Positive cases (n=373) |  |
| --- | --- | --- |
|  | n | % |
| Distribution of cases |  |  |
| Kars | 181 | 48.5 |
| Iğdır | 147 | 39.4 |
| Ardahan | 45 | 12.1 |
| Gender |  |  |
| Male | 156 | 41.8 |
| Female | 217 | 58.2 |
| Mean age | 36.96±20.25 |  |
| Median age | 33 |  |
| Age range |  |  |
| 0-20 | 70 | 18.8 |
| 21-40 | 152 | 40.8 |
| 41-60 | 108 | 28.9 |
| 60+ | 43 | 11.5 |
| Specimen types |  |  |
| Nasopharyngeal swab | 370 | 99.2 |
| Sputum | 0 | 0 |
| Tracheal aspirate | 3 | 0.8 |
| Symptoms |  |  |
| Cough | 111 | 29.8 |
| Fever | 98 | 26.3 |
| Sore throat | 49 | 13.2 |
| Shortness of breath | 43 | 11.5 |
|  | 40 | 10.7 |
| Headache | 38 | 10.2 |
| Myalgia | 18 | 4.8 |
| Stomachache | 7 | 1.9 |
| Diarrhea | 6 | 1.6 |
| Vomiting | 6 | 1.6 |
| Asymptomatic | 198 | 53.1 |
| Additional diseases |  |  |
| Respiratory diseases | 29 | 7.8 |
| Hypertension | 17 | 4.6 |
| Cardiovascular diseases | 12 | 3.2 |
| Diabetes | 9 | 2.4 |
| Cancer | 4 | 1.1 |

## Discussion

To our knowledge, this is one of the first epidemiologic study about the RT-PCR positivity of SARS-CoV-2 suspected cases in our country. In 60 days, we studied 10,000 specimens to detect SARS-CoV-2; however, the screening tests were excluded (n=2147). The positivity rates may vary depending on the study population and location. A study reported that 38% of 4880 were detected positive by RT-PCR [4]. Similarly, another study reported that the positivity rate was 51% [7]. The positivity rate of our population was 4.75%, which is almost compatible with the national positivity rates announced every day by the Minister of Health [8].

Pan et al. performed an epidemiologic study with 32.583 laboratory confirmed COVID-19 cases and reported that 51.6% of cases were female [9]. The percentages of positive male and female are parallel with the current literature. Among the 3211 female and 4642 male cases, females had a higher rate (n=217, 6.75%) of confirmed cases compared to males (n=156, 3.36%). This is parallel with the current literature. Although the number of tested males were more than one and half times from the number of tested females, the positivity rate in female was nearly two-fold of male (6.75% vs 3.36%). In our study, most positive cases were in the 21-40 (40.8%) and 41-60 (28.9%) age ranges, respectively. Similarly, a study from China performed with 44.672 cases indicated that most positive cases were in the 30-79 (87%) age range [10]. The average range was generally above 55 years [7]; however, this data in our study was about 38 years. This might be due to inclusion the large number of asymptomatic and close contact patients who were younger in our study.

On the other hand, cough and fever were the most seen symptoms in positive cases, respectively; however, most of the cases were those who had close contact with a COVID-19 patient (asymptomatic patients). Most of the studies reported that the number of asymptomatic patients were high and the most common symptoms were fever and cough [3, 11, 12]. Interestingly, total positive percentage were associated with gender, but not age in our study. On the contrary, a study reported that SARS-CoV-2 infection was statistically related with both age and gender [4].

As a conclusion, the epidemiologic studies should be performed about the prevalence of SARS-CoV-2 infection to better understand the effect of the virus all over the world.

### Study Limitations

Our study have some limitations. First, this was a retrospective study. Second, more detailed information should have been obtained because there were relatively large amounts of missing data for some outcomes. Third, the pandemic is still ongoing and the data about SARS-CoV-2 positivity can be change; however, we wanted to share the suspected patients’ rates just before the screening test has been started in our country.

## Data Availability

the availability of all data referred to in the manuscript

## Acknowledgement

Thanks for the physicians and/or workers of Kars, Iğdır, and Ardahan Pandemic Hospitals for invaluable supports to the preparation and transportation of samples.

## References

1. Zhai P, Ding Y, Wu X, Long J, Zhong Y, Li Y. The epidemiology, diagnosis and treatment of COVID-19. Int J Antimicrob Agents doi:10.1016/j.ijantimicag.2020.105955 105955 (2020).

2. Zimmermann P, Curtis N. Coronavirus Infections in Children Including COVID-19: An Overview of the Epidemiology, Clinical Features, Diagnosis, Treatment and Prevention Options in Children. Pediatr Infect Dis J 39(5), 355–368 (2020).

3. Bi Q, Wu Y, Mei S et al. Epidemiology and transmission of COVID-19 in 391 cases and 1286 of their close contacts in Shenzhen, China: a retrospective cohort study. Lancet Infect Dis doi:10.1016/S1473-3099(20)30287-5 (2020).

4. Liu R, Han H, Liu F et al. Positive rate of RT-PCR detection of SARS-CoV-2 infection in 4880 cases from one hospital in Wuhan, China, from Jan to Feb 2020. Clin Chim Acta 505 172–175 (2020).

5. Fang Y, Zhang H, Xie J et al. Sensitivity of Chest CT for COVID-19: Comparison to RT-PCR. Radiology doi:10.1148/radiol.2020200432 200432 (2020).

6. Xie X, Zhong Z, Zhao W, Zheng C, Wang F, Liu J. Chest CT for Typical 2019-nCoV Pneumonia: Relationship to Negative RT-PCR Testing. Radiology doi:10.1148/radiol.2020200343200343 (2020).

7. Chen N, Zhou M, Dong X et al. Epidemiological and clinical characteristics of 99 cases of 2019 novel coronavirus pneumonia in Wuhan, China: a descriptive study. Lancet 395(10223), 507–513 (2020).

8. https://covid19.saglik.gov.tr/ (x30.05.2020).

9. Pan A, Liu L, Wang C et al. Association of Public Health Interventions With the Epidemiology of the COVID-19 Outbreak in Wuhan, China. JAMA doi:10.1001/jama.2020.6130 (2020).

10. Wu Z, Mcgoogan JM. Characteristics of and Important Lessons From the Coronavirus Disease 2019 (COVID-19) Outbreak in China: Summary of a Report of 72314 Cases From the Chinese Center for Disease Control and Prevention. JAMA doi:10.1001/jama.2020.2648 (2020).

11. Hu Z, Song C, Xu C et al. Clinical characteristics of 24 asymptomatic infections with COVID-19 screened among close contacts in Nanjing, China. Sci China Life Sci 63(5), 706–711 (2020).

12. Sun P, Qie S, Liu Z, Ren J, Li K, Xi J. Clinical characteristics of hospitalized patients with SARS-CoV-2 infection: A single arm meta-analysis. J Med Virol doi:10.1002/jmv.25735 (2020).

